# Revisiting minimally important changes for the Oxford Hip and Knee scores

**DOI:** 10.1101/2025.07.29.25332390

**Authors:** Adam B. Smith, Damian Lewis, Stuart Mealing, Andria Joseph

## Abstract

**Purpose:** A number of metrics and thresholds have been proposed to evaluate minimally meaningful change in the Oxford Hip and Oxford Knee scores (OHS and OKS). The study aim was to evaluate the impact of baseline scores on these metrics, and whether treatment success could be evaluated beyond these single metrics.

**Patients & Methods:** The data were collated from the National Health Service in England including the OHS, OKS and a global transition item (GTI). Minimally important change (MIC) scores were derived and categorised by the GTI, baseline and post-operative score categories. These metrics were also evaluated against standard error of measurement (SEM).

**Results:** A total of 387,524 records were extracted. Although the overall MIC were in-line with previous research, the results showed these measures varied by pre-operative scores. Few MIC estimates exceeded measurement error thresholds. Those that did were fell in the category of post-operative scores ≥ 30 for both instruments.

**Conclusions:** The MIC for both the OHS and OKS are potentially unreliable single metrics for evaluating meaningful change. A combination of baseline, post-intervention and change scores may provide more a robust measure for the evaluation of patient outcomes and healthcare services.

## Introduction

Patient-reported outcome (PRO) measures capture aspects of individuals’ health-related quality of life (HRQoL) directly from the patient [1] and are important in helping to determine what matters most to patients in the management of their health. The Oxford Hip (OHS) and Oxford Knee Scores (OKS) [2,3] were developed to measure function and pain following hip and knee replacement. These measures have been used extensively in clinical research, have demonstrated robust psychometric properties [4]. and are routinely employed in the UK National Health Service (NHS) to evaluate patient outcomes and quality of care [5].

An important aspect of this evaluation is interpreting the meaningfulness of change in PRO scores. Metrics such as the minimally important change (MIC) may help in this process. Although different definitions abound[6], central to the MIC concept is that it reflects the smallest beneficial change perceived by the individual [7].

A number of studies have derived MIC and MID estimates for the OKS and OHS [8,9]. These range from around 9 to 12 for the OHS depending on whether used for evaluating primary hip replacement or revision surgery, and for the OKS between 9 and 11 [7,8]. More recent research has developed categories for interpreting clinically meaningful change in the OKS [10] with change scores ≥ 16 indicating significant improvement (“much better”) in patients’ subjective rating of their health and scores between 7 and 15 a moderate improvement (“little better”). This study also determined higher boundary scores dependent on baseline scores with the thresholds between categories increasing to ≥ 18 and 9-17 (thresholds between, respectively, “a little” / “much better”, and “about the same” / “a little better”) for those patients with baseline scores less than the mean OKS (<19).

The latter study indicates that baseline scores may impact on the MIC and potentially also the MID. In addition to these change metrics, attempts have also been made to define post-operative success either in terms of score categories, e.g., “excellent” or “good” outcome [11,12,13] or score thresholds beyond which surgical outcomes could be defined as successful [14,15,16]. Composite score thresholds including multiple patient-reported evaluations of surgical success have been shown to vary with baseline scores [15]. However, these factors aside, little or no attention has been focused on whether the combination of final outcome could be included alongside the baseline score and degree of change metrics to aide in the interpretation of meaningful change.

The purpose of this study was therefore to further elaborate meaningful change associated with the Oxford Hip and Oxford Knee scores by combining baseline and final scores with the minimally important change (MIC).

## Methods

### Instruments and Data

The Oxford Hip and the Oxford Knee Scores (OHS; OKS) [2,3] are two 12-item. Individual items for these PROs are reverse scored from 4 (“None” or “No”) to 0 (“Severe”, “Impossible”, “Unbearable”). A higher score reflects better function (maximum score: 48).

The data were collated from the publicly available data published between 2015 to 2022[5]. Patients undergoing either hip or knee surgery complete the OHS or OKS prior to and 6 months after surgery, in addition to rating post-surgery improvement in their hip/knee on a 5-point global transition item (“much better”; “a little better”; “about the same”; “a little worse”; and “much worse”).

### Statistical Analysis

Following previous research [8] the minimally important change (MIC) was derived using the mean change from baseline for those patients reporting “a little better” on the global transition item. The minimal important difference (MID) was calculated as the mean difference in change from baseline scores between patients reporting “about the same” and those who were “a little better”.

The degree of measurement error of a PROM may be estimated using the standard error of measurement (SEM). Some authors have proposed that the MIC should be greater than 4 times the SEM in order to distinguish the MIC metric from measurement error [17]. The 4SEM criterion was calculated using data from those patients recording no change in their health (“about the same”) [17] using the standard deviation of change (SD_change_) x √(1-Internal Reliability). Internal reliability was derived from Cronbach’s alpha for the baseline responses and was 0.89 for both instruments.

Both baseline, pre-operative scores as well as post-operative scores for the two PROMs were defined as follows using a combination of previously published categories [11] and thresholds [14,16]: scores ≤ 19; ≥20 and ≤ 29; ≥ 30 and ≤ 39; and >40.

The MIC was derived and summarised (mean and SD) by the global transition as well as baseline and post-surgery categories. In addition to this change scores were summarised by baseline score for both instruments and evaluated against the respective 4SEM criteria. The analysis was undertaken using R version 4.4.2.

## Results

A total of 184,509 patients completed the OHS; 203,015 had completed the OKS. The mean baseline score for OHS was 18 (Standard deviation, SD: 8.32) and 40 (SD: 8.57) at 6 months. For the OKS these were 19.28 (7.76) and 36.23 (9.34). The SEM for the OHS was 2.96 (4SEM, 11.85) and 2.42 (4SEM, 9.68) for the OKS.

The overall change from baseline was 22.01 (SD 10.13) and 16.95 (9.78) for the OHS and OKS, respectively (Table 1). Both instruments showed an increase in change scores or improvement across the global transition item from “much worse” to “much better”.

**Table 1.** Mean change from baseline in Oxford Hip & Knee Scores by baseline categories and global transition item.

| **Oxford Hip Score**<br>**Baseline /**<br>**GTI** | **N** | **Overall** | **Much worse** | **Little worse** | **About the same** | **MIC / Little better** | **Much better** |
| --- | --- | --- | --- | --- | --- | --- | --- |
| **≤19** | 109,817 | 26.09<br>(9.36) | 3.2<br>(10.5) | 6.5 (7.7) | 10.5<br>(8.6) | 15.3<br>(7.6) | 28.1<br>(7.6) |
| **≥20 & ≤29** | 57,033 | 18.02<br>(7.25) | -1.8<br>(10.8) | 0.9 (7.3) | 5.6 (7.9) | 9.5 (6.7) | 19.5<br>(5.7) |
| **≥30 & ≤39** | 16,040 | 10.34<br>(6.28) | -7.6<br>(10.5) | -3.8<br>(7.1) | 1.0 (6.8) | 4.0 (6.2) | 11.7<br>(4.7) |
| **≥40** | 1,619 | 1.22<br>(6.49) | -20.2<br>(9.0) | -7.8<br>(7.3) | -3.1<br>(5.7) | -2.0<br>(6.9) | 2.9 (4.6) |
| **Total** | 184,509 | 22.01<br>(10.13) | 0.60<br>(11.34) | 3.13<br>(8.53) | 7.68<br>(8.93) | 12.56<br>(8.22) | 23.81<br>(8.88) |
| **Oxford Knee Score**<br>**Baseline /**<br>**GTI** | **N** | **Overall** | **Much worse** | **Little worse** | **About the same** | **Little better** | **Much better** |
| **≤19** | 106,656 | 20.42<br>(9.86) | 0.7 (7.4) | 5.0 (6.4) | 8.0 (6.9) | 13.4<br>(6.9) | 24.3<br>(7.3) |
| **≥20 & ≤29** | 75,501 | 14.60<br>(7.78) | -4.0<br>(8.7) | 0.6 (6.4) | 3.8 (6.5) | 8.4 (6.0) | 17.2<br>(5.7) |
| **≥30 & ≤39** | 19,723 | 8.12<br>(6.70) | -9.2<br>(8.3) | -3.5<br>(6.6) | -0.6<br>(6.2) | 3.1 (5.8) | 10.2<br>(4.8) |
| **≥40** | 1,135 | 0.99<br>(6.44) | -16.9<br>(10.5) | -8.5<br>(7.0) | -6.8<br>(6.4) | -3.1<br>(5.8) | 2.9 (4.3) |
| **Total** | 203,015 | 16.95<br>(9.78) | -1.49<br>(8.55) | 2.59<br>(7.05) | 5.85<br>(7.30) | 10.84<br>(7.35) | 20.00<br>(8.14) |

The overall MIC for OHS was 12.56 and 10.84 for the OKS. Both metrics therefore exceeded the 4SEM criterion. This was not the case when change scores were categorised by baseline scores. Just under 60% patients had baseline OHS scores ≤ 19. The MIC for these patients was 15.3, i.e., > than 4SEM. However, this metric decreased (from 9.5 to −2) as the patient baseline scores increased, meaning 4SEM criterion was only achieved (in addition to the patients ≤ 19 reporting a “little better”) for those patients with baseline scores ≤ 29 who reported “Much better” improvement (change scores 19.5 to 28.1).

The same pattern was observed for OKS where 53% of patients had baseline scores ≤ 19: the MIC was 13.4 (decreasing from 8.4 to −3.1 for the remaining baseline categories). Again, only those patients reporting “much better” improvement had change scores exceeding the 4SEM criterion.

The results for the baseline analysis showed for the OKS, that for baseline scores ≤ 20, the change score for those patients reporting “a little better” health (i.e., the MIC), their mean change score fell between 11 and 16 (Figure 1). The range for the associated follow-up scores fell between 20 and 28. As the mean change scores decreased with increasing baseline scores, the change dropped below the 4SEM criterion (∼10) for baseline scores >20. Beyond this point any change score for patients reporting “a little better” would potentially not be distinguishable from measurement error. Similarly, patients’ baseline scores ≤ 20 for those “much better” were associated with a change score ranging between 20 and 29 (follow-up score range ≥ 40); the change score ranged between 10 and 20 (follow-up ≥ 30) for these patients with baseline scores greater than 20. For those patients reporting “about the same” level of health or no change in their status, the change scores ranged ± 10, i.e. with ± 4SEM, and change scores falling within this range may therefore not be discernible from measurement error. By extension, this applies to all change scores within this range irrespective of patients’ – if symmetry holds between magnitude of deterioration and improvement – for instance, those patients whose health was a “little worse” their change scores fell largely within this range (± 4SEM). For patients reporting “much worse” health, only those with baseline scores ≥40 reported change scores >-10.

**Figure 1:**
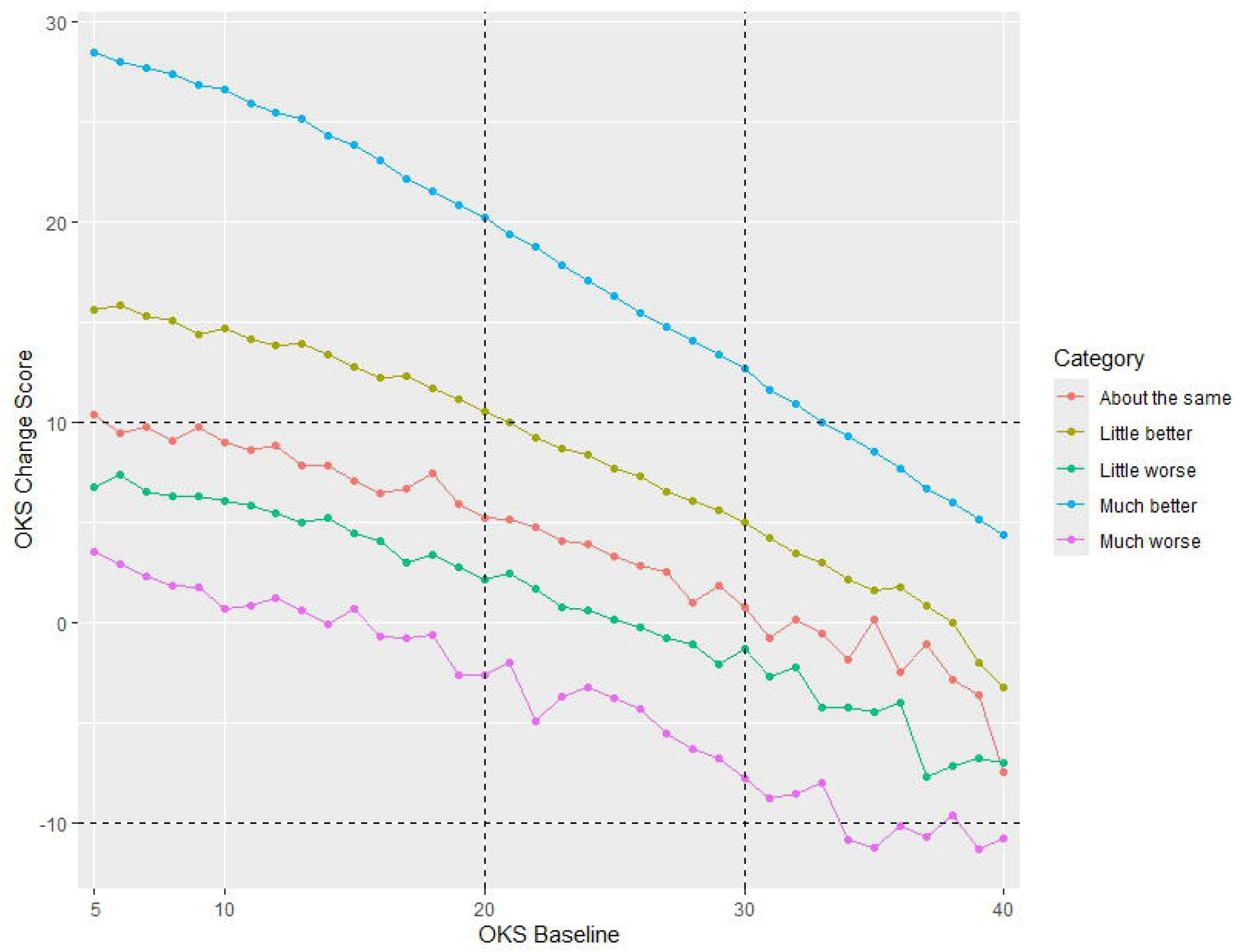

The OHS baseline analysis is summarised in Figure 2. Change scores for the “little better” group fell between 13 and 18 (MIC). These patients’ baseline scores were <20 and follow-up scores fell between 20 and 29. For those reporting “much better” health, their change scores fell between 20 and 29 with follow-up score ≥ 40. The majority of patients’ reporting no change in their health status had change scores within the ±4SEM criterion (assuming as with the OKS, symmetry between improvement and deterioration), and could potentially be construed as measurement error, although a few patients’ change scores were ≥ 13. The results for those patients reporting deterioration in their health status were less clearcut but suggested that patients with change score <-12 whose baseline scores were >40 reported their health as a “little worse”, whilst baseline scores >30 with the same change score were “much worse”.

**Figure 2:**
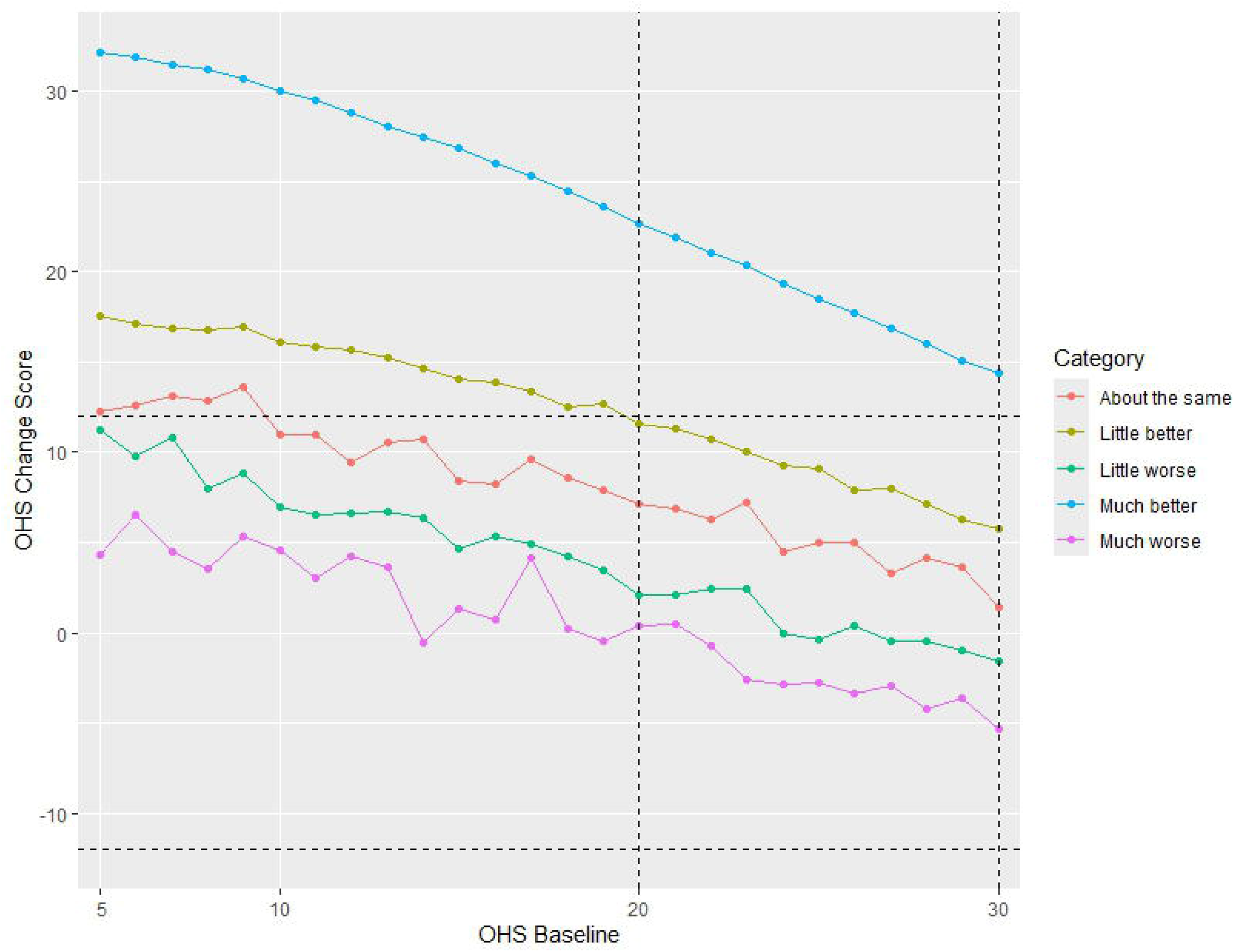

## Discussion

The aim of this study was to determine whether a combination of metrics including the baseline scores, post-operative scores and change scores could be utilised to identify meaningful change in both the OHS and OKS to aide interpretation of patient outcomes.

The results for the overall MIC were in-line with previous research [8,9] which had shown that baseline / pre-operative scores may affect the level of MIC [10]. This was confirmed in the current study: the magnitude of the MIC changed in line with baseline scores, decreasing as pre-operative score increased for both PROs.

However, significantly, the results further demonstrated that only those patients with baseline scores ≤ 20 recorded an MIC that exceeded the measurement error threshold (4SEM) for both PROMs.

The more nuanced analysis of baseline scores also revealed that only those change scores within the two categories of improvement (“Little better” / “Much better”) fell beyond the limits of measurement error. Change scores for the other three categories for both instruments were within measurement error, meaning deterioration in scores on these two instruments are difficult to interpret. Although, the assumption has been made here that measurement error was symmetrical, in other words that the 4SEM criterion equally applies to negative change.

Furthermore, the results also demonstrated, for those change scores >4SEM, that the MIC (and the other change categories), in combination with baseline OHS and OKS scores, was better characterised as a range, rather than single metric. For the OKS, scores between 7 and 15 have been suggested as indicating a “little better”. It is evident from the current study that a degree of these change scores (between 7 and 10) would potentially fall within the boundaries of measurement error (±10) [10].

These authors have proposed change scores ≥ 16 as indicators of patients faring “much better” [10]. The results of this study demonstrated that change scores ≥20 with baseline scores ≤ 20 are more likely to reflect a more robust indicator of a significant improvement in patient health.

Additionally, thresholds (≥ 30) have been proposed in the literature for evaluating treatment success for these PROMs [13,14,15]. The results of this study suggest that post-operative scores ≥ 20 with baseline scores ≤ 20 and change scores within the range 10-16 (OKS) or 12-18 (OHS) may represent a little improvement in patients’ health status on the OKS. Post-operative scores ≥30 and ≥ 40 may reflect significant improvement dependent on the baseline scores (Supplementary Table 1).

The main study limitation is the fact that no direct interpretation of patient-reported change in health was available with the analysis solely reliant on a single item response. Nevertheless, this should be balanced in mitigation against the large sample sizes.

In summary, the interpretation of meaningful change, specifically improvement, on both the OHS and OKS is dependent on three factors, namely the magnitude of change, and both the baseline and the post-operative scores. Although the anchor-based approach using a single metric with the global transition item is the preferred method[1] for deriving the MIC these results indicate that reliance on a single MIC alone to interpret meaningful change is inadequate in the context of these two PROMs.

## Conclusion

The MIC for both the OHS and OKS is a potentially an unreliable guide as a single metric for evaluating meaningful change. A combination of baseline, post-intervention and change scores may provide more a robust measure for the evaluation of patient outcomes and healthcare services.

## Data Availability

All data produced are available online at:
https://digital.nhs.uk/data-and-information/data-tools-and-services/data-services/patient-reported-outcome-measures-proms#latest-provisional-data

## Acknowledgements

Not applicable

