## Supplementary table for "Revisiting minimally important changes for the Oxford Hip and Knee scores"

**Supplementary Table 1. Outcomes Summary**

| **Oxford Hip Score -Baseline scores** | **Change score** | **Outcome category** | **Range follow-up scores** |
| --- | --- | --- | --- |
| < 20 | > 12 and ≤ 18 | Little better / MIC | ≥ 20 to < 30 |
| < 20 | ≥ 20 and ≤ 29 | Much better | ≥ 40 |
| <20 | ± 12 | Measurement error |  |
| ≥25 and ≤ 35 | >12 and ≤ 18 | Much better | ≥ 40 |
| >30 | <-12 | Deterioration Much worse | <0 |
| **Oxford Knee Score – Baseline scores** | **Change score** | **Outcome category** | **Range follow-up scores** |
| ≤ 20 | > 10 and ≤ 16 | Little better / MIC | ≥ 20 to ≤ 28 |
| ≤ 20 | ≥ 20 and ≤ 29 | Much better | ≥ 40 |
| Any | ≥ -10 and ≤10 | Measurement error | ≤ 16 |
| >20 | ≤10 | Measurement error | Any |
| >20 | > 10 and <20 | Much better | ≥ 30 |

1. Oxford Hip Score: Between -8 and 10 covers the range of those patients whose health was “about the same”, **plus 4SEM** is 10 for the OHS, therefore scores within this range fall within “measurement error”.
2. Knee Score: Between 0 and 12 covers the range of those patients whose health was “about the same”, **plus 4SEM** is 12 for the OKS, therefore scores within this range fall within “measurement error”.
